# Effect of Advanced Primary Care on Total Cost of Care

**DOI:** 10.64898/2026.08.21.26360946

**Authors:** Sasha Brodsky, Olga S. Matlin

**Author notes:** Corresponding author: Sasha Brodsky.

## Abstract

Improving primary care is a long-standing strategy to constrain health care spending. Yet, evaluations of primary care models focused on payment reform have shown minimal effects on total cost of care. We report the results from a large-scale, real-world evaluation of an advanced primary care model that restructures access through same-day and next-day appointments, on-demand video visits, asynchronous clinician messaging, and extended hours. Using a stacked-cohort difference-in-differences design with entropy balancing and inverse probability of censoring weighting, we analyzed multi-payer claims covering April 2022 through March 2025. Advanced primary care use was associated with an 8.6% reduction in total cost of care (-$729 per patient per year; P = 0.004), driven by lower specialist cost (-$939/year; P < 0.001) and, to a lesser degree, by reductions in inpatient (-$134/year; P < 0.001), urgent care (-$70/year; P < 0.001), and emergency department cost (-$16/year; P = 0.02), partially offset by higher primary care cost (+$350/year; P < 0.001). The specialist reduction was concentrated in knowledge-based consultative encounters (-$663/year; P < 0.001), while procedural specialist cost was largely unchanged (-$276/year; P = 0.09). Cost differences emerged in the first post-index month. These findings suggest that advanced primary care may reduce total health care spending, with observed savings driven primarily by lower spending on consultative specialty care.

## 1. Introduction

In 2024, US health care spending reached nearly 2.5 times the OECD average on a per capita basis.^1^ Yet, less than 5% of US health care spending is devoted to primary care, compared with 14% in other high-income countries.^2^ As the entry point to the health care system and the setting for longitudinal care and care coordination, primary care has been central to policy efforts aimed at improving value and moderating spending growth.^2,3^

Efforts to improve primary care have not consistently reduced total health care spending. Under the Comprehensive Primary Care Plus (CPC+) model, enhanced payments combined with requirements for care delivery transformation did not reduce total expenditures among Medicare^4^ or commercially insured beneficiaries.^5^ Evaluations of patient-centered medical homes, which reorganized primary care around enhanced access, care coordination, and population management, have found heterogeneous and generally modest effects on spending.^6^ Expanding access through new sites or modalities has not lowered spending either: retail clinics and direct to consumer telehealth increased spending because most encounters represented new use rather than substitution for more costly care.^7,8^ This evidence suggests that incremental additions to conventional primary care may be insufficient to meaningfully reduce spending.

We evaluated One Medical, an advanced primary care model, that combines same-day and next-day appointment availability, on-demand video visits, asynchronous clinician messaging that supports ongoing symptom evaluation and medication management, and extended hours access. Patients access it through either employer-sponsored membership or direct to consumer enrollment. We do not observe enrollment channel in the data, and our estimates capture the combined effect of the model across both pathways. Patients in both channels face standard health plan cost sharing, including applicable copayments, deductibles, and coinsurance. The model therefore expands access to primary care without introducing differential financial incentives that could independently affect utilization. We evaluated the effect of this advanced primary care model on total health care spending.

## 2. Methods

### 2.1 Data Source

We used the McKesson Compile Data platform, a multi-payer data resource that aggregates adjudicated medical and pharmacy claims for commercially insured and Medicare (fee-for-service and Medicare Advantage) beneficiaries across the United States. Individuals are represented by a de-identified, tokenized patient identifier consistent across claims, enabling longitudinal follow-up within and across payers. The study period spanned April 1, 2022 through March 31, 2025, encompassing 12 months of baseline observation, the 12-month period during which patients could enter the treatment cohort (April 2023–March 2024), and 12 months of post-index follow-up. The study was reviewed by the WCG Institutional Review Board and determined to be exempt from further review as secondary analysis of existing de-identified data. We followed the RECORD (Reporting of Studies Conducted Using Observational Routinely Collected Health Data) extension of the STROBE statement.

### 2.2 Study Population

We defined 12 monthly cohorts covering April 2023 through March 2024. For each cohort, the index month was the calendar month of the patient’s qualifying primary care visit, and each patient contributed 12 baseline and 12 post-period months relative to their own index month. Primary care visits were defined by provider specialty (family medicine, internal medicine, general practice, geriatric medicine, nurse practitioner, or physician assistant) and place of service (office, telehealth, federally qualified health center, rural health clinic, or independent clinic).

Treatment group included patients whose index month contained a primary care visit at the advanced practice, identified by matching the rendering clinician’s National Provider Identifier (NPI) to the practice’s clinician roster as of the visit date. Comparison patients were those whose index month contained a primary care visit at any other provider within the same metropolitan statistical area (MSA). Both groups were required to have no visit to the advanced practice in the 12 months preceding their index month. Additional eligibility criteria applied to both groups: at least one claim in both the baseline and post periods, at least one primary care visit in the index month, annualized total allowed costs below $200,000 in the baseline period, and no baseline claims related to oncology, dialysis, end-stage renal disease, transplant, hospice, inpatient rehabilitation, or specialty infusion therapy. From this eligible pool we drew a comparison sample without replacement at a 50:1 ratio, stratified by MSA and cohort month.^1^ A comparison patient meeting eligibility criteria in more than one cohort could be sampled multiple times, contributing a patient-cohort record with its own index month. Applying these criteria yielded 109,809 treated and 1,826,386 comparison patient-cohort records, each observed for 12 months before and after the index visit. Because allowed cost amounts were not observed in both the pre-and post periods for every record, the analytic sample was restricted to 25,052 treated and 752,400 comparison patient-cohort records (**eTable 1**). We applied inverse probability of censoring weighting (IPCW), reweighting this sample to represent the full eligible population.

### 2.3 Outcomes

We defined total allowed cost as the sum of allowed amounts on medical claims and total paid amounts on pharmacy claims. Primary care, specialist, emergency department, inpatient, and urgent care costs were identified via place of service codes and CPT/HCPCS classification. We additionally examined the likelihood of any specialist cost and a partition of specialist cost into consultative and procedural claims. Consultative claims were defined as evaluation and management (E&M) visits and other knowledge-based specialist services that can be rendered through primary care. Procedural claims were defined as surgical, imaging, endoscopic, or similar services requiring specialist equipment or skill, identified using Current Procedural Terminology (CPT) and Healthcare Common Procedure Coding System (HCPCS) code ranges. Claims with any procedural line item were classified as procedural; all remaining specialist claims were classified as consultative.

### 2.4 Statistical Analysis

We used a stacked-cohort difference-in-differences (DiD) design.^9,10^ Because treated and comparison patients differed on observable baseline characteristics, we reweighted the comparison group using entropy balancing (EB) weights,^11^ fit on the pooled sample to match the treatment group on the first and second moments of baseline utilization by type (PCP, specialist, ED, inpatient, urgent care), age, age 65 indicator, sex, Charlson comorbidity index, frailty score, count of chronic conditions, high-utilizer flag (at least two ED visits or at least one hospitalization at baseline), and five chronic-condition indicators (diabetes, heart failure, COPD/asthma, hypertension, and menopause).

To correct for non-random selection into the cost observed sample, we introduced a second weighting step. IPCW weights were derived from a logistic regression of a complete cost record indicator on baseline covariates.^12^ The model was fit with five-fold cross-fitting: predicted probabilities used to construct each patient’s weight were derived from folds that excluded that patient, preventing own observation contamination (**eTable 2**). Combining EB and IPCW into a compound weight recovered an unbiased estimate of the average treatment effect on the treated (ATT) in the presence of both observed covariate imbalance and non-random cost observability.

The primary estimator was a patient-level weighted least squares regression of the within-patient change in outcome (the mean of the outcome over the 12 post-period months minus the mean over the 12 baseline months) on a treatment indicator, baseline covariates, and cohort-month fixed effects, weighted by the compound EB and IPCW. Cohort-month fixed effects absorbed differences in cohort composition and calendar time shifts. This first-difference specification is numerically equivalent to a stacked two-way fixed effects DiD on the collapsed pre-post panel, absorbing patient and calendar time fixed effects by construction. Statistical significance was defined as two-sided P < 0.05. Category-level cost outcomes are reported as descriptive decompositions of the primary total cost of care estimate, and no formal correction was applied given the pre-specified hierarchical structure.

The two groups differed before weighting (**Table 1**). Treated patients were younger (mean age 42.4 vs. 51.0 years), more evenly split by sex (52.4% female vs. 61.5%), and carried a lower comorbidity burden (mean chronic conditions 0.64 vs. 2.83; 11.1% classified as high utilizers vs. 35.1%) with lower baseline utilization across every category we examined. EB drove all standardized mean differences (SMDs) to below 0.001 on the cost observed sample. Compound weights, which correct for differential attrition into cost observability, retained a maximum SMD of 0.086 (baseline specialist visits/month). The second largest was 0.075 (MSK/chronic pain), with all remaining covariates having SMD below 0.08.

**Table 1.** Covariate Balance Before and After Weighting.

| Covariate | Treated | Comp:<br>Raw | Comp:<br>EB | Comp:<br>EB×IPCW | \|SMD\|<br>Raw | \|SMD\|<br>EB | \|SMD\|<br>EB×IPCW |
| --- | --- | --- | --- | --- | --- | --- | --- |
| **Demographics** |  |  |  |  |  |  |  |
| Age (years) | 42.39 | 51.04 | 41.72 | 41.20 | 0.520 | 0.040 | 0.072 |
| Age 18-34 | 0.364 | 0.227 | 0.364 | 0.380 | 0.304 | <0.001 | 0.036 |
| Age 35-44 | 0.298 | 0.160 | 0.298 | 0.290 | 0.334 | <0.001 | 0.019 |
| Age 45-54 | 0.140 | 0.166 | 0.140 | 0.140 | 0.073 | <0.001 | 0.001 |
| Age 55-64 | 0.074 | 0.185 | 0.074 | 0.075 | 0.335 | <0.001 | 0.003 |
| Age ≥ 65 | 0.123 | 0.262 | 0.123 | 0.114 | 0.356 | <0.001 | 0.023 |
| Female | 0.524 | 0.615 | 0.524 | 0.506 | 0.184 | <0.001 | 0.036 |
| Male | 0.476 | 0.385 | 0.476 | 0.494 | 0.184 | <0.001 | 0.036 |
| **Comorbidity** |  |  |  |  |  |  |  |
| Charlson index | 0.182 | 0.852 | 0.182 | 0.163 | 0.589 | <0.001 | 0.017 |
| Chronic conditions | 0.637 | 2.828 | 0.637 | 0.574 | 1.206 | <0.001 | 0.034 |
| High utilization | 0.111 | 0.351 | 0.111 | 0.109 | 0.593 | <0.001 | 0.007 |
| **Chronic Condition Type** |  |  |  |  |  |  |  |
| Diabetes | 0.028 | 0.183 | 0.028 | 0.025 | 0.520 | <0.001 | 0.012 |
| Heart failure | 0.005 | 0.031 | 0.005 | 0.004 | 0.192 | <0.001 | 0.009 |
| Hypertension | 0.054 | 0.353 | 0.054 | 0.045 | 0.801 | <0.001 | 0.022 |
| Hyperlipidemia | 0.051 | 0.401 | 0.051 | 0.045 | 0.921 | <0.001 | 0.016 |
| Coronary artery disease | 0.013 | 0.067 | 0.013 | 0.010 | 0.278 | <0.001 | 0.015 |
| Chronic kidney disease | 0.009 | 0.055 | 0.009 | 0.007 | 0.266 | <0.001 | 0.011 |
| COPD/asthma | 0.043 | 0.196 | 0.043 | 0.038 | 0.484 | <0.001 | 0.018 |
| Obesity | 0.029 | 0.200 | 0.029 | 0.025 | 0.558 | <0.001 | 0.011 |
| Mental-health disorders | 0.119 | 0.362 | 0.119 | 0.111 | 0.592 | <0.001 | 0.020 |
| MSK/chronic pain | 0.198 | 0.545 | 0.198 | 0.186 | 0.769 | <0.001 | 0.027 |
| GERD/GI | 0.066 | 0.307 | 0.066 | 0.059 | 0.651 | <0.001 | 0.018 |
| Allergic rhinitis | 0.021 | 0.128 | 0.021 | 0.019 | 0.417 | <0.001 | 0.009 |
| Peri-/post-menopause | 0.002 | 0.017 | 0.002 | 0.002 | 0.149 | <0.001 | 0.002 |
| **Baseline Utilization** |  |  |  |  |  |  |  |
| PCP visits/month | 0.094 | 0.431 | 0.094 | 0.092 | 0.568 | <0.001 | 0.003 |
| Specialist visits/month | 0.258 | 0.472 | 0.258 | 0.210 | 0.281 | <0.001 | 0.063 |
| ED visits/month | 0.014 | 0.041 | 0.014 | 0.010 | 0.248 | <0.001 | 0.039 |
| Hospitalizations/month | 0.008 | 0.020 | 0.008 | 0.006 | 0.104 | <0.001 | 0.016 |
| Urgent-care visits/month | 0.012 | 0.018 | 0.012 | 0.012 | 0.102 | <0.001 | 0.006 |
| **Payer Mix** |  |  |  |  |  |  |  |
| Commercial | 0.856 | 0.637 | 0.856 | 0.872 | 0.520 | <0.001 | 0.040 |
| Medicare | 0.115 | 0.215 | 0.115 | 0.094 | 0.272 | <0.001 | 0.058 |
| Medicaid | 0.013 | 0.121 | 0.013 | 0.014 | 0.442 | <0.001 | 0.006 |
| VA / DoD / Federal | 0.008 | 0.014 | 0.008 | 0.007 | 0.054 | <0.001 | 0.008 |
| Other / Unknown | 0.008 | 0.013 | 0.008 | 0.012 | 0.049 | <0.001 | 0.038 |

EB = entropy balancing; IPCW = inverse probability of censoring weights; SMD = standardized mean difference; Comp = comparison group.

To trace the treatment effect across the 12 pre-index and 12 post-index months, we estimated an event-study model on the patient-month panel using the same compound weights as the DiD. The regressors were patient-cohort fixed effects, event-time indicators for each month relative to the index month, and interactions between event-time and the treatment indicator, with month −1 as the omitted reference category. Standard errors were cluster-robust at the patient level to account for within-patient serial correlation across months.^13^ Month 0, the calendar month containing the patient’s qualifying primary care visit, was excluded from both the DiD and event study specifications because it contains the initial exposure. The same specification served two purposes: (1) joint F-tests of the 11 baseline treatment and event-time coefficients provided a parallel-trends check (excluding month -1 as a reference month); and (2) post-period coefficients traced the monthly evolution of the treatment effect.

To address potential anticipation in the immediate pre-index months, we fit a donut specification excluding the three months preceding the index month.^14^ Nine of the ten cost outcomes satisfied parallel trends in the standard specification, and urgent care satisfied parallel trends in the donut specification (**eTable 3**). As a further falsification check, we estimated a within-baseline placebo by splitting the baseline in half and treating event-time t = −6 as a placebo index: months [−12, −7] served as the placebo baseline and months [−6, −1] as the placebo follow-up. Placebo coefficients were statistically indistinguishable from zero for eight of the ten cost outcomes, and the two exceptions moved in the direction opposite to the main effects (**eTable 4**).

## 3. Results

Advanced primary care reduced total cost of care by $729 per patient per year (8.6% relative to treatment group’s baseline mean; P = 0.004). The largest reduction was in specialist cost (-$939/year; 20.7%; P < 0.001), followed by inpatient (-$134/year; P < 0.001), urgent care (-$70/year; P < 0.001), and ED (-$16/year; P = 0.02). Primary care costs increased by $350/year (P < 0.001), offsetting 30% of the $1,159/year reductions across specialist, ED, inpatient, and urgent care. Pharmacy cost was unchanged (+$10/year; P = 0.86). Total medical cost (excluding pharmacy) decreased by $739/year (P = 0.003). Full monthly and annual DiD coefficients for all outcomes are reported in **eTable 5**.

Differences in cost emerged in the first post-index month with no gradual buildup (**Figure 2**). The effect of advanced primary care on specialist, ED, and urgent care costs turned negative in month 1 and remained so through month 12. Baseline coefficients clustered around zero across all cost outcomes except urgent care, consistent with the parallel trends assumption for the cost outcomes that drive the main result. The temporal pattern, with specialist cost divergence beginning in the first post-index month and no gradual buildup, is consistent with shifts in care-seeking behavior at the point of entry rather than slower acting mechanisms such as chronic disease management.

To characterize the mechanism behind the reduction in specialist cost, we examined two outcomes: whether patients had any specialist contact, and how cost changed among patients with any specialist care (**Figure 3**). Advanced primary care reduced the likelihood of any specialist contact by 9.85 percentage points (-23.4% relative reduction from a 42.1% treatment group baseline; P < 0.001). This reduction was concentrated in consultative care, where contact fell 10.04 percentage points (-29.6% from a 34.0% baseline; P < 0.001), compared with a smaller decline in procedural contact (-3.26 percentage points or -15.2% from a 21.4% baseline; P < 0.001). Unconditional specialist cost estimates aligned directionally with this pattern: cost reductions were concentrated in consultative claims (−$663/year; P < 0.001) and were not statistically distinguishable from zero for procedural claims (−$276/year; P = 0.09).

The results were robust to alternative specifications. Only one of the ten cost outcomes had a placebo coefficient statistically distinguishable from zero, and it moved in the direction opposite to the main effects (**eTable 4**). After adjusting for the baseline placebo differential, the point estimates were larger in magnitude than the main results, though wider confidence intervals rendered the adjusted total cost, specialist cost, and consultative cost estimates non-significant at conventional levels: −$788/year for total cost of care (P = 0.25, vs. main −$729), −$1,041/year for specialist cost (P = 0.11), and −$699/year for consultative specialist cost (P = 0.19). Adjusted estimates remained statistically significant for inpatient (−$141/year, P = 0.005) and urgent care (−$109/year, P = 0.005), the two categories most likely to reflect concurrent access-driven substitution. Results were also robust to using EB weights alone, which yielded a total cost of care effect of −$653/year (**eTable 6**).

## 4. Discussion

Advanced primary care reduced total cost of care by $729/year within 12 months, primarily through specialist cost substitution. The magnitude and speed of the reduction contrasts with the null or modest findings of prior primary care interventions. Payment reform CPC+ model produced no total cost of care reduction in either Medicare^4^ or commercial^5^ populations. Medical home transformations produced modest single-digit reductions in utilization but inconsistent effects on total costs.^6^ Access expanding retail-clinic and telehealth interventions increased condition-level spending by generating new utilization rather than substituting for higher cost visits.^7,8^ These models modified payment or access in isolation. In contrast, advanced primary care restructures access as part of an integrated primary care model.

Reductions in specialist costs accounted for most of the reduction in total cost of care. The reduction emerged in the first post-index month and persisted through month 12, a pattern more consistent with early-stage shifts in care-seeking than with slower disease management or hospitalization prevention mechanisms. The reduction was concentrated in consultative specialist claims (−$663/year; P < 0.001) rather than procedural claims (−$276/year; P = 0.09), consistent with primary care substituting for E&M functions that do not require specialist-specific equipment or skill. The likelihood of any specialist contact was 9.85 percentage points lower in the advanced primary care group (P < 0.001), further indicating that the reduction operated at the point of care initiation.

### 4.1 Limitations

Several considerations bear on interpretation. First, entropy balancing corrects imbalance on observed characteristics but cannot rule out unobserved determinants of practice choice such as patient motivation or health-seeking behavior. Placebo analysis showing no baseline divergence constrains the plausible magnitude but does not eliminate this concern. Second, cost outcomes are unobserved for a subset of claimants. IPCW addressed this and estimates were insensitive to its inclusion (−$653/year without IPCW vs. −$729 with). Third, the 12-month follow-up may not fully distinguish specialist care that was averted from specialist care that was deferred, and longer horizons are needed to observe downstream effects of substituted care. Fourth, claims data do not reveal whether reductions reflect clinician gatekeeping, patient-initiated substitution, or improved condition management. Fifth, both study arms are care-seeking by construction (defined by a primary care visit in the index month), and findings apply to commercially insured adults with claims-based access to primary care and may not generalize to Medicare, Medicaid, uninsured, or rural populations.

### 4.2 Policy Implications

For payers and employers, including advanced primary care options may yield savings when access incentives target members with specialist-heavy utilization. For policymakers, the results imply that delivery redesign focused on access may complement payment reform models. As CMS continues to expand value-based purchasing, pairing payment reform with access restructuring could accelerate the shift from episodic specialty care to longitudinal primary care management that neither reform achieves in isolation.

## 5. Conclusion

Initiating care at an advanced primary care practice was associated with an 8.6% reduction in total cost of care within 12 months of index, driven predominantly by lower specialist cost. The immediacy of the reduction and its concentration in consultative specialty care are consistent with primary care substituting for specialty visits at the point of care-seeking, rather than with slower disease management or hospitalization prevention pathways.

## 8. Acknowledgments

The authors thank David Card, PhD, Susan Murphy, PhD, and Jessie Handbury, PhD, for critical review of the manuscript, and Trupti Yenpreddiwar for data construction. None received compensation for their contributions outside of their usual compensation. The authors acknowledge using large language model-based AI tools to help draft and refine the manuscript text based on author-conceived ideas and outlines. Specifically, we used Anthropic Claude Opus 4.7 accessed through Amazon Bedrock via the Cline IDE assistant. All intellectual content, including study conception, design, analysis, interpretation, and conclusions, originated from the authors. All AI-generated output was critically reviewed, edited for accuracy, and approved by the authors, who take full responsibility for the content of this manuscript.

## 9. Ethics Statement

This work is a retrospective analysis of secondary de-identified administrative claims data licensed from a third-party health data platform (McKesson Compile Data). Because the analysis used pre-existing, de-identified administrative data with no direct patient contact, no study-specific intervention, and no re-identification of individuals, it qualifies as non-human-subjects research under 45 CFR 46.102 and did not require institutional review board approval.

## 10. Data availability

The claims data underlying this study were licensed by Amazon from McKesson Compile Data and contain de-identified administrative records subject to a commercial data use agreement. Redistribution of the underlying patient-level records is prohibited by that agreement and by contractual restrictions with the participating payers. Investigators seeking direct access to the underlying claims data should contact McKesson Compile Data directly through their standard licensing pathway.

## 11. Code availability

The statistical analysis was performed in Python 3.12 using open-source libraries (pandas, numpy, statsmodels, scikit-learn, matplotlib).

## Online Supplement

### Effect of Advanced Primary Care on Total Cost of Care

This supplement accompanies the main manuscript and provides additional methodological detail, sensitivity analyses, and tabular results. All estimates derive from the same analytic file as the main manuscript.

We plot monthly cost level for the treated cohort and the EB and IPCW-reweighted comparison cohort across event-time months. Baseline levels are visually parallel across arms, and the mechanical index-month spike appears in every panel, most visibly for Urgent Care because care delivered in the index month is correlated with the reason for seeking primary care. Index month is excluded from the DiD. Post-period divergence between arms is concentrated in Specialist Cost and its consultative sub-component, mirroring the DiDs reported in **Figure 1**.

**Figure 1.**
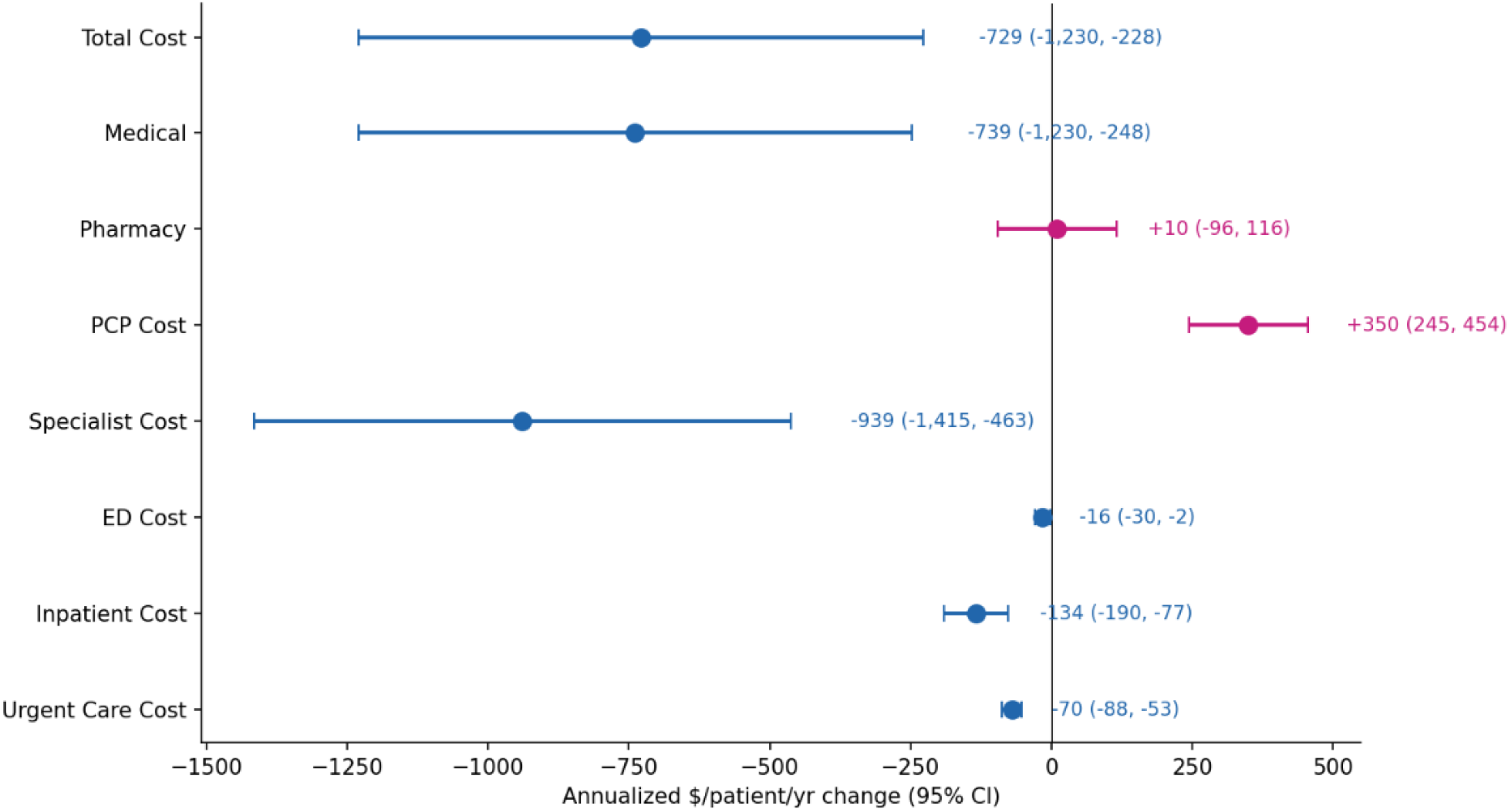
Effect of Advanced Primary Care on Cost by Category Note: Point estimates and 95% confidence intervals for the DiD effect of initiating care at an advanced primary care practice on annualized allowed cost per patient by cost category. Estimates are from patient-level weighted least squares regression on the within-patient change from pre- to post-index period, weighted by compound EB and IPCW, adjusted for baseline covariates and cohort-month fixed effects. Sample includes 25,052 treated and 752,400 comparison patient-cohort records. Standard errors are cluster-robust at the patient level. Positive values indicate higher cost among advanced primary care patients. Negative values indicate lower cost.

**Figure 2.**
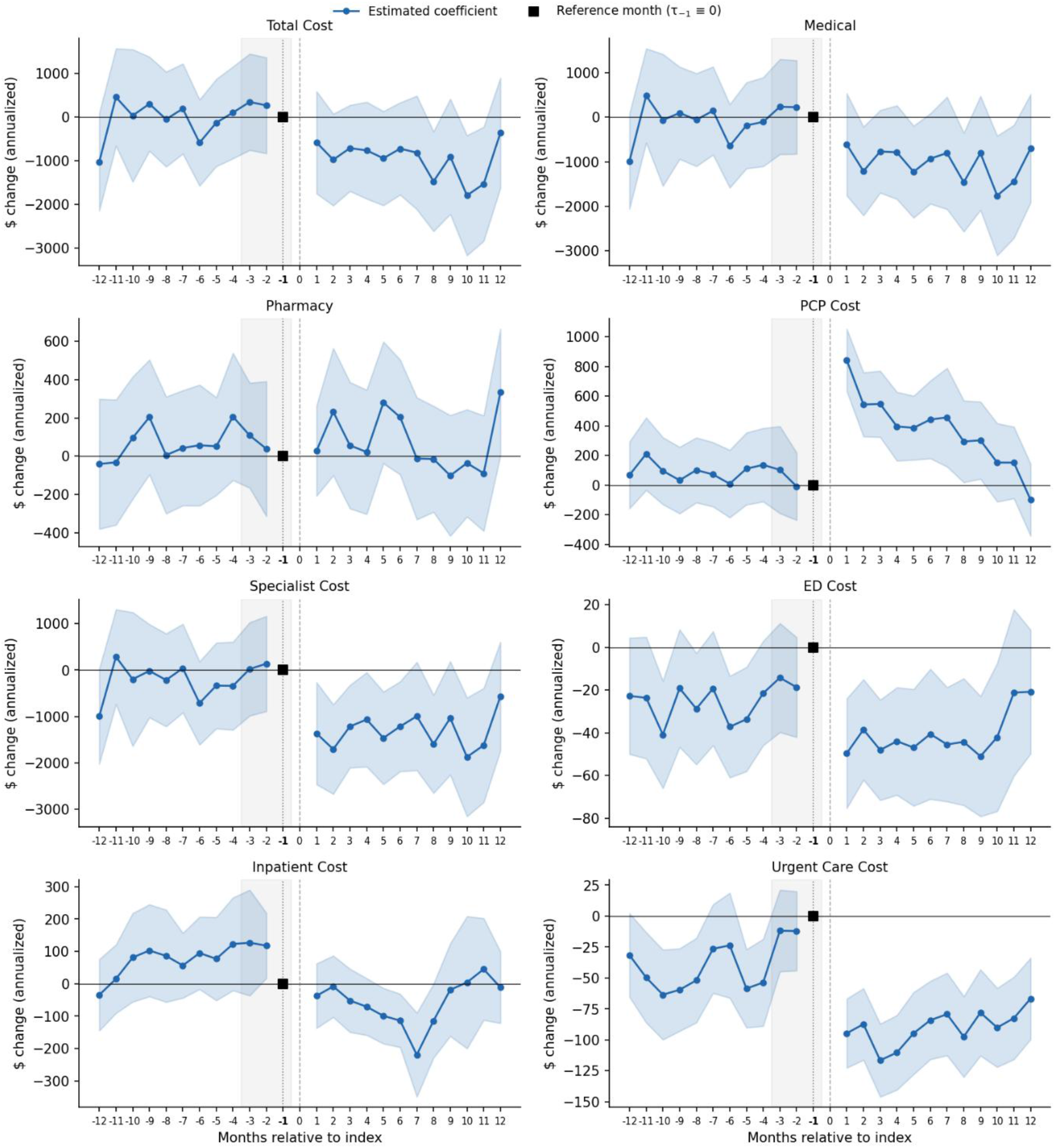
Event Study of Cost by Category Note: Monthly treatment and event-time interaction coefficients from an event study specification fit on the patient-month panel using compound EB and IPCW, with patient-cohort fixed effects. Coefficients represent the difference in monthly allowed cost per patient between advanced primary care and comparison groups, relative to month −1 (omitted reference). Month 0 (the index month) is excluded from both pre- and post-periods. Sample includes 25,052 treated and 752,400 comparison patient-cohort records. Error bars represent 95% confidence intervals. Standard errors are cluster-robust at the patient level. Parallel-trends F-tests on the 11 pre-period coefficients: Total Cost F = 0.86, P = 0.58; Specialist Cost F = 0.69, P = 0.74.

**Figure 3.**
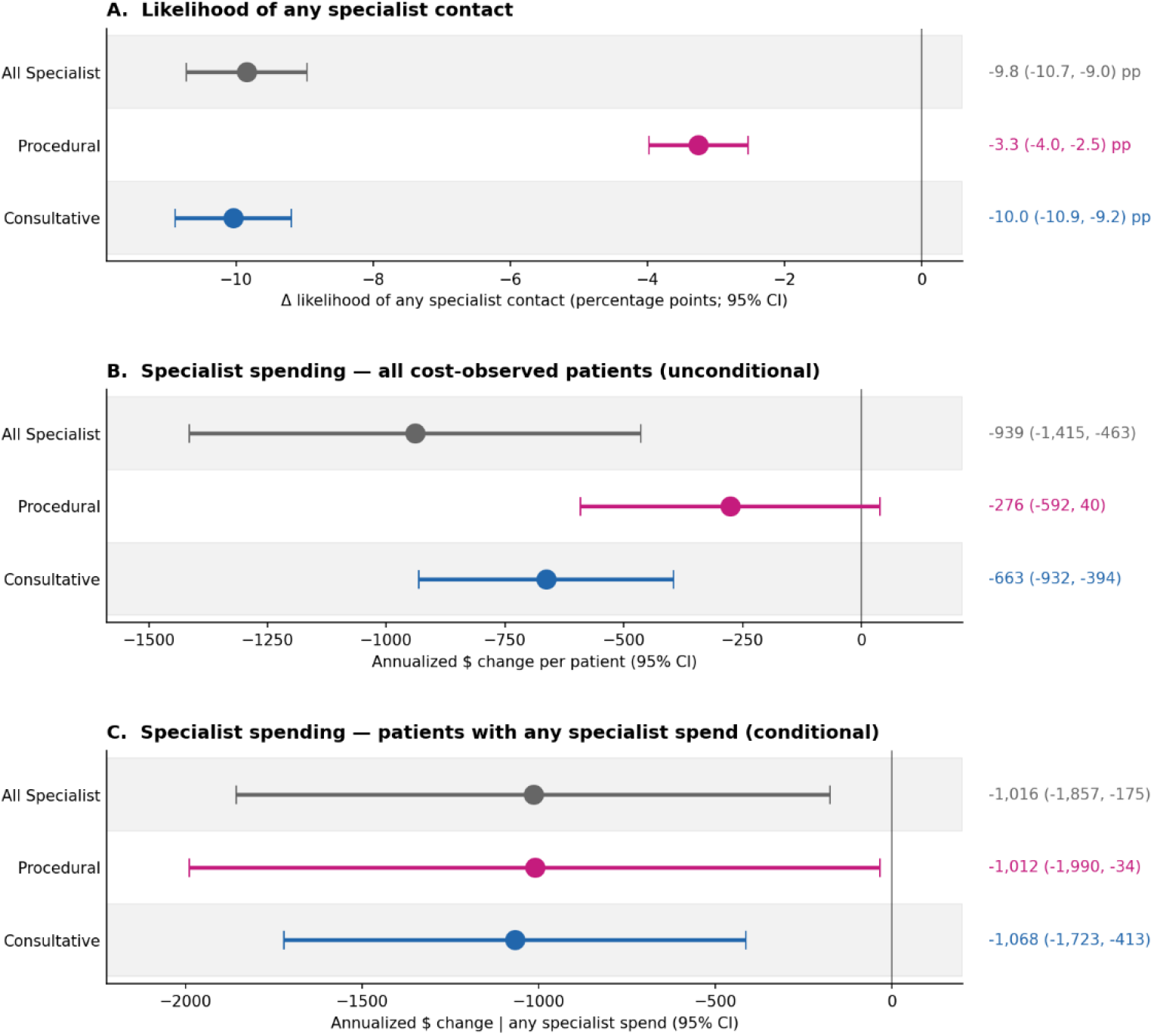
Effect of Advanced Primary Care on Consultative vs Procedural Specialist Cost Note: Panel A, change in likelihood of any specialist contact (percentage points); Panel B, unconditional specialist cost ($/patient/year); Panel C, conditional specialist cost among patients with any specialist contact ($/patient/year). DiD estimates from patient-level weighted least squares regression, weighted by compound EB and IPCW, adjusted for baseline covariates and cohort-month fixed effects. Panel A reports the change in likelihood of any specialist contact (percentage points). Panel B is the effect on specialist cost per patient per year among all patients (including those with no specialist contact). Panel C is the effect on specialist cost per patient per year among the subset of patients with any specialist contact. Consultative claims are specialist claims with no procedural CPT/HCPCS code. Procedural claims include surgical, imaging, infusion, endoscopic, or device-implantation codes. Sample includes 25,052 treated and 752,400 comparison patient-cohort records. Error bars represent 95% confidence intervals. Standard errors are cluster-robust at the patient level. Statistical significance was defined as P < 0.05.

**eFigure 1.**
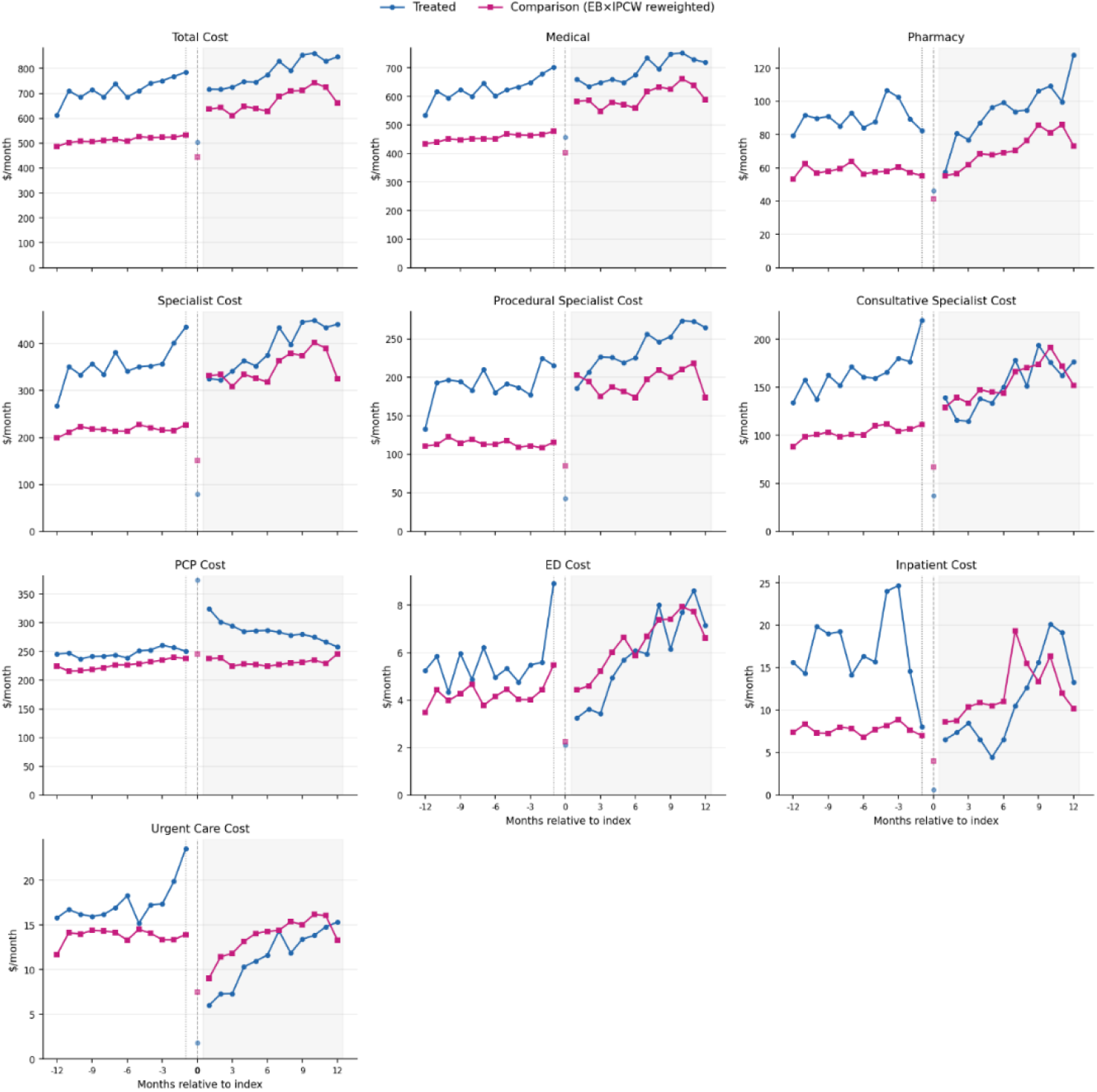
Level Differences across Treatment and Comparison Groups. Note: Total allowed cost per patient is defined as the sum of allowed amounts across all medical and pharmacy claims. Allowed amount reflects negotiated reimbursement and is unaffected by patient cost-sharing structure. Other outcomes follow the same definition restricted to claims of a particular care setting using rendering provider taxonomy code and place of service code. We decompose specialist cost into procedural and consultative components. A specialist claim is classified as procedural if any line item carries a CPT/HCPCS code in the procedural code set (surgical, imaging, infusion, endoscopic, or device-implantation codes); otherwise the claim is classified as consultative.

Sample construction begins with the MSA-stratified pool of 680,845 treatment-eligible patients and 5,237,463 matched comparison patients and excludes patients without a first eligible primary care visit and ≥12 months of pre/post primary care use, those with annualized total allowed costs > $200,000 in the baseline period, and those with no allowed amounts.

**eTable 1.** Sample Construction Funnel.

| Step | Description | Treated N | Comparison N |
| --- | --- | --- | --- |
| 1 | Patients identified (50:1 MSA-stratified comparison pool) | 680,845 | 5,237,463 |
| 2 | First eligible primary care visit in Apr 2023–Mar 2024 | 115,319 | 5,237,463 |
| 3 | Excluding patients with annualized total allowed costs $> \$200,000$ | 109,809 | 1,826,386 |
| 4 | Patients with any observed costs in baseline and follow-up periods | 25,052 | 752,400 |

We report diagnostics for the cross-fit logistic IPCW censoring model. We fit the model on 1,406,940 patients, estimating each patient’s probability of cost observability. The Brier score of 0.248 is at marginal baseline of 0.247, indicating IPCW covariates carry little additional predictive information about cost observability. IPCW adjustment corrects from small differences in observability trends across arms, as evidenced by differential attrition test. Hosmer-Lemeshow, reported for completeness, is not informative in this sample size.

**eTable 2.** IPCW Censoring Model Diagnostics.

| Diagnostic | Value |
| --- | --- |
| Observations used to fit calibration | 1,406,940 |
| Observations in $M = 1$ (cost-observed) subsample | 777,452 |
| Brier score | 0.248 |
| Hosmer-Lemeshow $\chi^2$ | 114,403.81 |
| Hosmer-Lemeshow P | $< 0.001$ |
| Differential-attrition test, interaction term | -0.43 |
| Differential-attrition test, interaction term P | $< 0.001$ |

Notes:
- Brier score is the mean squared error between predicted probabilities of cost observability and the observed outcome. It ranges from 0 (perfect prediction) to 1 (worst possible prediction). The marginal/baseline score is obtained by predicting the overall observed-cost rate for every patient, serving as a reference point. - Hosmer-Lemeshow test groups patients into deciles of predicted probability and compares observed versus predicted event rates within each group - A differential attrition test is a linear probability specification of the observability indicator on the interaction of treatment and post period indicators, adjusted for covariates and clustered at the patient level.

Joint F-tests of the 11 baseline treatment and event-time interactions from the event-study specification (event-time −1 omitted as the reference) support the parallel trends assumption for every cost outcome that drives the results. Of the 10 cost outcomes, nine are consistent with parallel trends at the 5% level under the full specification (P ≥ 0.06). Urgent Care fails the full-specification test (P = 0.010) but passes the donut specification (P = 0.76), a pattern characteristic of anticipation effect in the immediate pre-index window.

**eTable 3.** Parallel Trends F-Tests: Full and Donut Specifications.

| **Outcome** | **Full P** | **Donut P (d=3)** |
| --- | --- | --- |
| Total Cost | 0.58 | 0.40 |
| Total Medical | 0.64 | 0.43 |
| Total Pharmacy | 0.95 | 0.92 |
| Primary Care | 0.88 | 0.86 |
| Specialist | 0.74 | 0.55 |
| Specialist, Procedural | 0.58 | 0.67 |
| Specialist, Consultative | 0.93 | 0.25 |
| Emergency Dept | 0.22 | 0.11 |
| Inpatient | 0.46 | 0.45 |
| Urgent Care | 0.010 | 0.76 |

The 12-month baseline is split in half, and event-time −6 is relabeled as a placebo treatment onset, with months [−12, −7] treated as the placebo baseline and months [−6, −1] as the placebo post period. Across the 10 cost outcomes the placebo coefficients are small and statistically indistinguishable from zero at conventional levels except for Urgent Care (+$39/year, P < 0.001), which moves in the direction opposite to the main effect. Placebo-adjusted point estimates are larger in magnitude than the main effects: Total Cost adjusted −$788/year (P = 0.25 vs. main −$729/year, P = 0.004), Specialist Cost adjusted −$1,041/year (P = 0.11 vs. main −$939/year, P < 0.001), Consultative Specialist Cost adjusted −$699/year (P = 0.19 vs. main −$663/year, P < 0.001), Inpatient adjusted −$141/year (P = 0.005 vs. main −$134/year, P < 0.001), and Urgent Care adjusted −$109/year (P = 0.005 vs. main −$70/year, P < 0.001). Adjusted estimates for Total Cost, Specialist Cost, and Consultative Specialist Cost are non-significant at conventional levels; adjusted estimates for Inpatient and Urgent Care remain statistically significant.

**eTable 4.** Baseline Placebo and Placebo-Adjusted DiD.

| **Outcome** | **Main $\beta$** | **Main P** | **Placebo $\beta$** | **Placebo P** | **Placebo-adj $\beta$** | **Adj. P** |
| --- | --- | --- | --- | --- | --- | --- |
| Total Cost | -729 | 0.004 | +58 | 0.83 | -788 | 0.25 |
| Total Medical | -739 | 0.003 | +35 | 0.90 | -774 | 0.17 |
| Total Pharmacy | +10 | 0.86 | +24 | 0.73 | -14 | 0.72 |
| Primary Care | +350 | <0.001 | -80 | 0.25 | +430 | 0.03 |
| Specialist | -939 | <0.001 | +102 | 0.69 | -1,041 | 0.11 |
| Specialist, Procedural | -276 | 0.09 | +66 | 0.70 | -342 | 0.22 |
| Specialist, Consultative | -663 | <0.001 | +36 | 0.80 | -699 | 0.19 |
| Emergency Dept | -16 | 0.02 | +6 | 0.42 | -22 | 0.19 |
| Inpatient | -134 | <0.001 | +7 | 0.83 | -141 | 0.005 |
| Urgent Care | -70 | <0.001 | +39 | <0.001 | -109 | 0.005 |

Baseline placebo analysis following Roth (2022). Months $[-12, -7]$ as placebo baseline, months $[-6, -1]$ as placebo post, fake event at $t = -6$ . Placebo coefficients are small and statistically indistinguishable from zero at conventional levels for every cost outcome except Urgent Care ( $P < 0.001$ ). Reported $\beta$ is in $\$/\text{yr}$ .

We report monthly DiD estimates for all 10 cost outcomes, with annualized effects shown for comparison with the manuscript. Advanced primary care reduces total cost of care by −$729/year (P = 0.004) and specialist cost by −$939/year (P < 0.001), driven by −$663/year on the consultative care (P < 0.001). Acute care cost falls in every category (ED −$16/year P = 0.02; Inpatient −$134/year P < 0.001; urgent care −$70/year P < 0.001), and primary care cost rises by +$350/year (P < 0.001). Total pharmacy cost is indistinguishable from zero (+$10/year, P = 0.86).

**eTable 5.** Main DiD Estimates using Compound Weights.

| Outcome | $\beta$ /month | SE | P | 95% CI | $\beta$/year |
| --- | --- | --- | --- | --- | --- |
| Total Cost | -60.77 | 21.30 | 0.004 | (-102.52, -19.02) | -729 |
| Total Medical | -61.60 | 20.87 | 0.003 | (-102.49, -20.70) | -739 |
| Total Pharmacy | +0.82 | 4.51 | 0.86 | (-8.02, +9.67) | +10 |
| Primary Care | +29.13 | 4.46 | <0.001 | (+20.39, +37.87) | +350 |
| Specialist | -78.26 | 20.22 | <0.001 | (-117.90, -38.62) | -939 |
| Specialist, Procedural | -23.01 | 13.42 | 0.09 | (-49.32, +3.30) | -276 |
| Specialist, Consultative | -55.25 | 11.42 | <0.001 | (-77.63, -32.86) | -663 |
| Emergency Dept | -1.32 | 0.59 | 0.02 | (-2.48, -0.17) | -16 |
| Inpatient | -11.15 | 2.40 | <0.001 | (-15.84, -6.45) | -134 |
| Urgent Care | -5.86 | 0.73 | <0.001 | (-7.30, -4.42) | -70 |

We compare the main effect to an EB-only specification using the same sample of 777,452 patients with observed cost. For total cost of care, primary care, specialist care and its sub-components, inpatient care, and urgent care, adding IPCW moves the point estimate by less than 10% and preserves statistical significance. Large shifts in ED cost and pharmacy cost reflect movement on outcomes whose baseline effect sizes are small.

**eTable 6.** Compound Weight (EB and IPCW) vs EB weight.

| Outcome | Main $\beta$ | Main P | EB $\beta$ | EB P | $\Delta$ |
| --- | --- | --- | --- | --- | --- |
| Total Cost | -729 | 0.004 | -653 | 0.01 | +11.7% |
| Total Medical | -739 | 0.003 | -639 | 0.01 | +15.6% |
| Total Pharmacy | +10 | 0.86 | -13 | 0.80 | -26.4% (ns) |
| Primary Care | +350 | <0.001 | +307 | <0.001 | +13.9% |
| Specialist | -939 | <0.001 | -804 | 0.002 | +16.9% |
| Specialist, Procedural | -276 | 0.09 | -196 | 0.19 | +40.7% (ns) |
| Specialist, Consultative | -663 | <0.001 | -607 | <0.001 | +9.1% |
| Emergency Dept | -16 | 0.02 | -15 | 0.008 | +5.5% |
| Inpatient | -134 | <0.001 | -127 | <0.001 | +5.0% |
| Urgent Care | -70 | <0.001 | -67 | <0.001 | +5.3% |

$\Delta$ = percent change in absolute magnitude of the IPCW estimate relative to EB. Both columns derive from the same 777,452 patient-level observations. IPCW-headline runs larger in magnitude than EB-only on Total, Medical, Specialist, Consultative, Procedural, Plan Paid, and Patient OOP outcomes; the conclusions are unchanged under either weighting choice. Reported $\beta$ is in \$/yr.

## Footnotes

1 MSAs with fewer than 20 times the treated count within each monthly cohort contributed their full eligible pool rather than a 50:1 sample.

